# Characteristics associated with culture-positivity versus antibiotic administration in patients with cirrhosis on ICU admission: a retrospective cohort study in 5 hospitals

**DOI:** 10.64898/2026.09.22.26363729

**Authors:** Andrew J. Davis, Chad H. Hochberg, Khyzer B. Aziz, Li Yan, Sarina Sahetya, David N. Hager, Theodore J. Iwashyna

**Author notes:** **CORRESPONDING AUTHOR:** Andrew J. Davis, Johns Hopkins University, 1830 E Monument Street, 5th Floor, Baltimore, MD 21287.

## Abstract

**Objective:** To test the hypothesis that selected SIRS and SOFA criteria measured at ICU admission have divergent associations with (1) the development of new culture-confirmed infections and (2) early empiric broad-spectrum antibiotic receipt in critically ill patients with cirrhosis.

**Design:** Retrospective, observational study using electronic health record (EHR) data.

**Setting:** Both community and academic ICUs within a five-hospital health system between July 2017 and December 2025.

**Subjects:** Adult patients >18 years of age diagnosed with cirrhosis admitted to the ICU within 96 hours of hospital admission. Patients transferred from outside facilities and those with positive cultures collected >24 hours before ICU admission were excluded.

**Interventions:** None.

**Measurements:** Multivariable logistic regression tested the association of the most aberrant values in nine SIRS/SOFA vital sign and laboratory parameters measured during the 24 hours prior to ICU admission with (1) the development a newly positive culture collected during the 24 hours before through the 96 hours after ICU admission and (2) receipt of broad-spectrum antibiotics during the 24 hours before through the 24 hours after ICU admission.

**Main Results:** The presence of hyperthermia, hypothermia, hypoxia, or hypotension at ICU admission was associated with both culture positivity and antibiotic receipt. The presence of leukocytosis was associated with antibiotic receipt but not culture positivity. Among patients in the 21.6% of encounters with newly positive cultures, normothermia and normal WBC were common (respectively present in 97.2% and 77.5% of encounters) and associated with antibiotic omission at ICU admission despite the subsequent development culture positivity.

**Conclusions:** Traditional physiologic markers of infection are often absent in critically ill patients with cirrhosis who develop new culture-confirmed infections at ICU admission, possibly contributing to delayed antibiotics. There may be opportunities to improve antibiotic delivery and stewardship at ICU admission in this high-risk population.

## INTRODUCTION

Critically ill patients with cirrhosis experience poor outcomes with 40-50% ICU mortality.^1,2^ Infections often drive decompensation and death in this population,^3-5^ as patients with cirrhosis are at increased risk of infections— both bacterial and fungal—due to cirrhosis-associated immune dysfunction.^6,7^ Occult infections frequently occur concurrently with non-infectious decompensations in this population; for example, approximately half of patients with cirrhosis and severe gastrointestinal bleeding also have an active infection.^8^ This presents a clinical challenge as delays in antibiotic administration—which greatly increase the risk of extrahepatic organ failure and death—may be more common in this population due to the presence of concomitant non-infectious critical illness syndromes.^2,9^

Diagnosing infections in this population may be challenging due cirrhosis-induced changes in physiology that confound and mimic traditional physiologic markers of infection.^3,4,10^ These changes include hyperdynamic circulation with hypotension and tachycardia,^11-13^ resting dyspnea and tachypnea,^14,15^ and impaired lactate clearance.^4,10,16^ Additionally, cirrhosis causes immune dysfunction that blunts fevers and leukemoid reactions.^4,17^ Some have even argued that patients with cirrhosis and sepsis may represent a unique phenotype of sepsis with distinct biomarker signatures.^18^ To balance the risks of missed infections with antimicrobial stewardship,^19,20^ critically ill patients with cirrhosis may require alternative strategies for the early detection of sepsis and administration of empiric broad spectrum antibiotics.^9,10,21-23^

We hypothesized that traditional vital sign and laboratory measurements commonly used to diagnosis infections and sepsis at ICU admission^24-28^—namely the systemic inflammatory response syndrome (SIRS) and sequential organ failure assessment (SOFA) criteria—would be (1) weakly associated with the development of new culture-confirmed infections (2) yet strongly associated early empiric broad-spectrum antibiotic receipt in critically ill patients with cirrhosis.

## METHODS

### Study Design, Setting, and Included Population

We performed a retrospective cohort study of all adults >18 years of age with cirrhosis admitted to an intensive care unit (ICU) in any of five hospitals (2 academic, 3 community) within the Johns Hopkins Health System (JHHS) between July and 2017 and December 2025 who were not known to have an active infection at the time of ICU admission (**Figure 1**).

**Figure 1:**
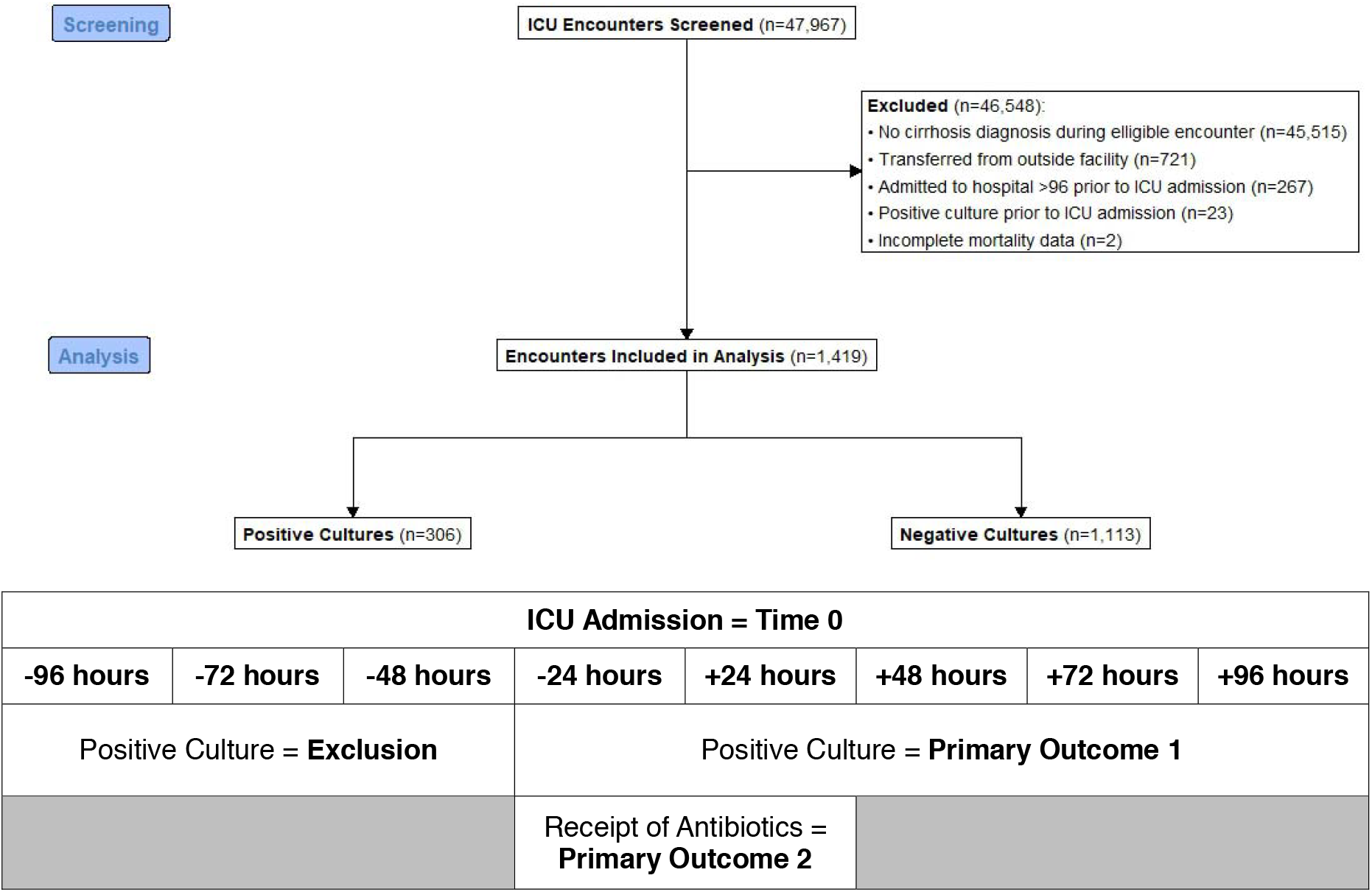
Consort Diagram and Study Design.

Cirrhosis diagnoses were defined by ICD-10 codes previously validated for use with electronic health record (EHR) data (supplementary **eTable 1**).^29^ Patients were assessed for eligibility at ICU admission and included in the analysis if their ICU admission was within 96 hours of hospital admission. Patients were eligible for reenrollment during subsequent hospital encounters after discharge (with appropriate adjustment for non-independence as outlined in the statistical methods).

Patients with positive cultures collected in the 24-96 hours prior to ICU admission were excluded. Patients who were transferred from outside hospitals or facilities were additionally excluded, as data for cultures collected prior to their ICU admission would not be reflected in our EHR.

### Data Source & Collection

Data were abstracted from the Johns Hopkins Critical Illness and Recovery Center of Excellence registry, which contains detailed clinical and operational EHR data on all JHHS encounters for patients admitted to any ICU throughout the JHHS. Abstracted data included age, sex, race, Elixhauser comorbidity index, hospital length of stay, and overall mortality (all demographic data were complete for included patients).^30^

After a review of the sepsis literature, data for a parsimonious list of physiologic parameters based on Sepsis-3 guidelines and elements of the SIRS and SOFA scores that are (a) most relevant to the diagnosis of infections, (b) commonly measured and resulted back to clinicians at the time of ICU admission, and (c) objectively quantifiable using EHR data were collected from the 24 hours prior to ICU admission.^27^ This included: maximum temperature (°C), minimum temperature (°C), maximum heart rate (beats/minute), minimum non-invasive mean arterial pressure (MAP, measured in mmHg), maximum respiratory rate (breaths/minute), minimum SpO_2_:F_I_O_2_ ratio,^31^ maximum white blood cell count (WBC, count/μL), minimum WBC, and maximum lactate (mmol/L).

The data were inspected for non-physiologic results consistent with errors, and nonsense values were censored. Missingness was assessed for each physiologic parameter, and missing values (including censored values) were imputed using multivariate imputation by chained equations with 10 replications.^32^ A summary of the physiologic data pre- and post-imputation is listed in supplementary **eTable 2** with list of censored values (13 values in total) in supplementary **eTable 3**.

Cultures obtained during the 96 hours before through 96 hours after ICU admission were followed until final results were available (i.e., results finalizing after 96 hours post-ICU admission were included). Culture sources included blood, ascites, other intraabdominal sources (e.g., biliary), genitourinary, cerebrospinal fluid (CSF), bronchoalveolar lavage (BAL), upper respiratory tract, and pleural fluid. Wound cultures, tissue cultures, and multi-drug-resistant screening swabs (e.g. nasopharyngeal and rectal screening swabs) were excluded. At the time these data were generated, no rapid nucleic amplification tests were available.^33^

### Outcomes

Primary outcomes were (1) the development of a newly positive culture collected (rather than resulted) during the 24 hours before through the 96 hours after ICU admission, and (2) receipt of broad-spectrum antibiotics during the 24 hours before through the 24 hours after ICU admission.

Cultures growing organisms inherently considered non-pathogenic from a given source (e.g., bronchoalveolar lavage with growth of candida) were considered negative (full list of culture results considered non-pathogenic in supplementary **eTable 4**).^34^ Cultures growing organisms sometimes attributed to possible contamination (e.g., blood culture growing coagulase negative staphylococcus) were considered positive given the high frequency of true pathogenic infections with such organisms in patients with cirrhosis.^35^ We excluded these cultures in a separate sensitivity analysis.

All eligible patients, regardless of whether a culture was collected during follow-up, were included in the analysis. While this may have introduced bias given that patients without cultures collected could not meet the culture positivity primary outcome (although they could still meet the antibiotic receipt primary outcome), our goal was to preserve an unselected study population in which there was diagnostic uncertainty as to whether an active infection was present.^36^ Limiting the analysis to patients that had cultures collected may introduce bias by selecting for a study population with a higher pre-test probability of having active infections present at the time of ICU admission. Therefore, in a separate sensitivity analysis we excluded patients who did not have any cultures collected during follow-up.

Patients were considered to have received broad-spectrum antibiotics if they received intravenous aztreonam, cefepime, daptomycin, ertapenem, linezolid, meropenem, piperacillin-tazobactam, or vancomycin—the most common broad-spectrum antibiotics in our system.^37^ Prophylactic antibiotics for gastrointestinal bleeding— ceftriaxone and ciprofloxacin—were not considered broad-spectrum antibiotics in this specific context as they are not recommended first-line antibiotics for critically ill patients requiring ICU admission within the studied health system (we recognize this may vary in other health systems).^38^ Secondary outcomes included 30-day mortality, 90-day mortality, and hospital length of stay.

In a “missed antibiotics” post-hoc analysis among patients with positive cultures, we further evaluated the association of the measured physiologic parameters with the odds of not receiving antibiotics during the 24 hours before through the 24 hours after ICU admission despite developing a positive culture.

### Statistical Analysis

Baseline characteristics of the cohort including demographics, physiologic data, and unadjusted outcome incidence were summarized using counts and proportions for categorical variables and medians and interquartile ranges (IQRs) for continuous variables. We then used multivariable logistic regression models to evaluate the association of the most aberrant value of each physiologic parameter measured during the 24 hours before ICU admission with each primary outcome separately. Models were adjusted for all collected physiologic parameters. To account for potential non-independence across multiple hospital encounters for the same patient, confidence intervals were computed using cluster-robust standard errors.^39^

Reported odds ratios were scaled to clinically meaningful intervals (e.g., odds of infection for a 5 beat per minute increase in maximum heart rate). To investigate if individual physiologic parameters had additive value in predicting the presence of culture-positive infections at ICU admission, we separately reported the area under the receiver operating characteristic (AUROC) for both the overall model adjusted for all measured physiologic parameters, as well as separate models with each individual physiologic parameter alone.

All analyses were performed using R version 4.5.2.^40^ Analytic code and log files are included as supplementary materials in the appendix.

## RESULTS

A total of 47,967 ICU encounters were screened for inclusion. After exclusions, 1,419 eligible ICU encounters—all from separate hospitalizations—comprising 1,249 unique patients were included in the final analysis (**Figure 1**). Baseline patient characteristics and clinical outcomes for all eligible encounters are reported in **Table 1**. Encounters were characterized by a patient cohort that had a median age of 59 (IQR 49-67) and was majority male (n=867, 61.1%), white (n=892, 62.9%), and non-Hispanic (n=1,156, 81.5%). ICD-10 codes for alcohol use disorder were present in approximately a quarter (n=379, 26.7%) of encounters.

**Table 1:** Baseline characteristics and outcome among all eligible ICU encounters stratified by culture positivity.

| Characteristic | Overall<br>N = 1,419<sup>1</sup> | Negative Cultures<br>N = 1,113<sup>1</sup> | Positive Cultures<br>N = 306<sup>1</sup> | p-value<sup>2</sup> |
| --- | --- | --- | --- | --- |
| Age | 59 (49, 67) | 59 (49, 67) | 59 (48, 67) | 0.9 |
| Female | 552 (39%) | 415 (37%) | 137 (45%) | **0.02** |
| Race |  |  |  | 0.3 |
| American Indian or Alaskan Native | 5 (0.4%) | 5 (0.4%) | 0 (0%) | - |
| Asian | 59 (4.2%) | 44 (4.0%) | 15 (4.9%) | - |
| Black or African American | 296 (20.8%) | 229 (20.6%) | 67 (21.9%) | - |
| Native Hawaiian or Pacific Islander | 6 (0.4%) | 6 (0.5%) | 0 (0%) | - |
| Other | 150 (10.6%) | 113 (10.2%) | 37 (12.1%) | - |
| White | 892 (62.9%) | 705 (63.3%) | 187 (61.1%) | - |
| Unknown | 11 (0.8%) | 11 (1.0%) | 0 (0%) | - |
| Ethnicity |  |  |  | 0.5 |
| Hispanic or Latino | 111 (7.8%) | 91 (8.2%) | 20 (6.5%) | - |
| Not Hispanic or Latino | 1,156 (81.5%) | 906 (81.4%) | 250 (81.7%) | - |
| Unknown | 152 (10.7%) | 116 (10.4%) | 36 (11.8%) | - |
| Alcohol Abuse | 379 (26.7%) | 289 (26.0%) | 90 (29.4%) | 0.2 |
| Obesity | 486 (34.2%) | 369 (33.2%) | 117 (38.2%) | **0.04** |
| Diabetes | 461 (32.5%) | 355 (31.9%) | 106 (34.6%) | 0.4 |
| Heart Failure | 386 (27.2%) | 290 (26.1%) | 96 (31.4%) | 0.06 |
| Chronic Pulmonary Disease | 328 (23.1%) | 246 (22.1%) | 82 (26.8%) | 0.08 |
| Received Antibiotics | 635 (44.7%) | 400 (35.9%) | 235 (76.8%) | **<0.001** |
| 30 Day Mortality | 252 (17.8%) | 168 (15.1%) | 84 (27.5%) | **<0.001** |
| 90 Day Mortality | 343 (24.2%) | 237 (21.3%) | 106 (34.6%) | **<0.001** |
| Length of Stay (Days) | 10.9 (5.8, 29.0) | 9.8 (5.3, 21.0) | 18.2 (9.6, 85.2) | **<0.001** |
| <sup>1</sup> Median (Q1, Q3); n (%), <sup>2</sup> Wilcoxon rank sum test, Pearson's Chi-squared test, or Fisher's exact test. |  |  |  |  |

Eligible cultures were collected in 64.3% (n=913) of encounters; positive cultures were detected in 21.6% (n=306) of encounters. Among the 306 encounters with positive cultures, 8.2% (n=25) had positive culture results comprised entirely of results considered potential contaminants. Compared to those without positive cultures, encounters with positive cultures had increased 30-day mortality (27.5% vs 15.1%), 90-day mortality (34.6% vs 21.3%), and hospital length of stay (18.2 days vs 9.8 days) (**Table 1**).

Among the 306 encounters with positive cultures, the most common sources of positive cultures were blood (56.2%, n=172), genitourinary (27.8%, n=85), upper respiratory (19.9%, n=61), and ascites (9.5%, n=29) with other sources accounting for 5.6% (n=17) of positive cultures (supplementary **eTable 5**). More than one source of positive cultures was observed in 16.0% (n=49) of encounters. Gram-positive and gram-negative organisms were respectively detected in 50.0% (n=153) and 50.7% (n=155) of encounters. Fungal organisms and mycobacterial organisms were respectively detected in 5.6% (n=17) and less than 1.0% (n=1) of encounters. Multiple types of organisms were found in 17.0% (n=52) of encounters. After excluding potential contaminant culture results in a sensitivity analysis, the distribution of positive culture sources was similar (supplementary **eTable 5**; see supplementary **eTable 6** for complete list of results characterized as potential contaminants).

### Parameters Associated with Positive ICU-Admission Cultures

Using multivariable logistic regression, hyperthermia (increases in maximum temperature), hypothermia (reductions in minimum temperature), hypoxia (reductions in minimum SpO_2_:F_I_O_2_ ratio), and hypotension (reductions in MAP) were significantly associated with culture positivity (**Figure 2**). The odds of culture positivity were greatest with hyperthermia (OR 1.61 per degree Celsius increase in maximum temperature, 95% CI 1.27-2.06) and hypothermia (OR 1.42 per degree Celsius decrease in minimum temperature, 95% CI 1.12-1.82). Including all physiologic measurements improved model discrimination for the detection of positive cultures at ICU admission (AUROC of 0.750) compared to any individual physiologic parameter alone (supplementary **eFigure 1**). The results of the sensitivity analysis excluding potential contaminant culture results were similar except that hypothermia was no longer associated with culture positivity (supplementary **eTable 7**). The results of the sensitivity analysis excluding patients without cultures collected during follow-up were also similar except that maximal respiratory rate was now associated with culture positivity (supplementary **eTable 8**).

**Figure 2:**
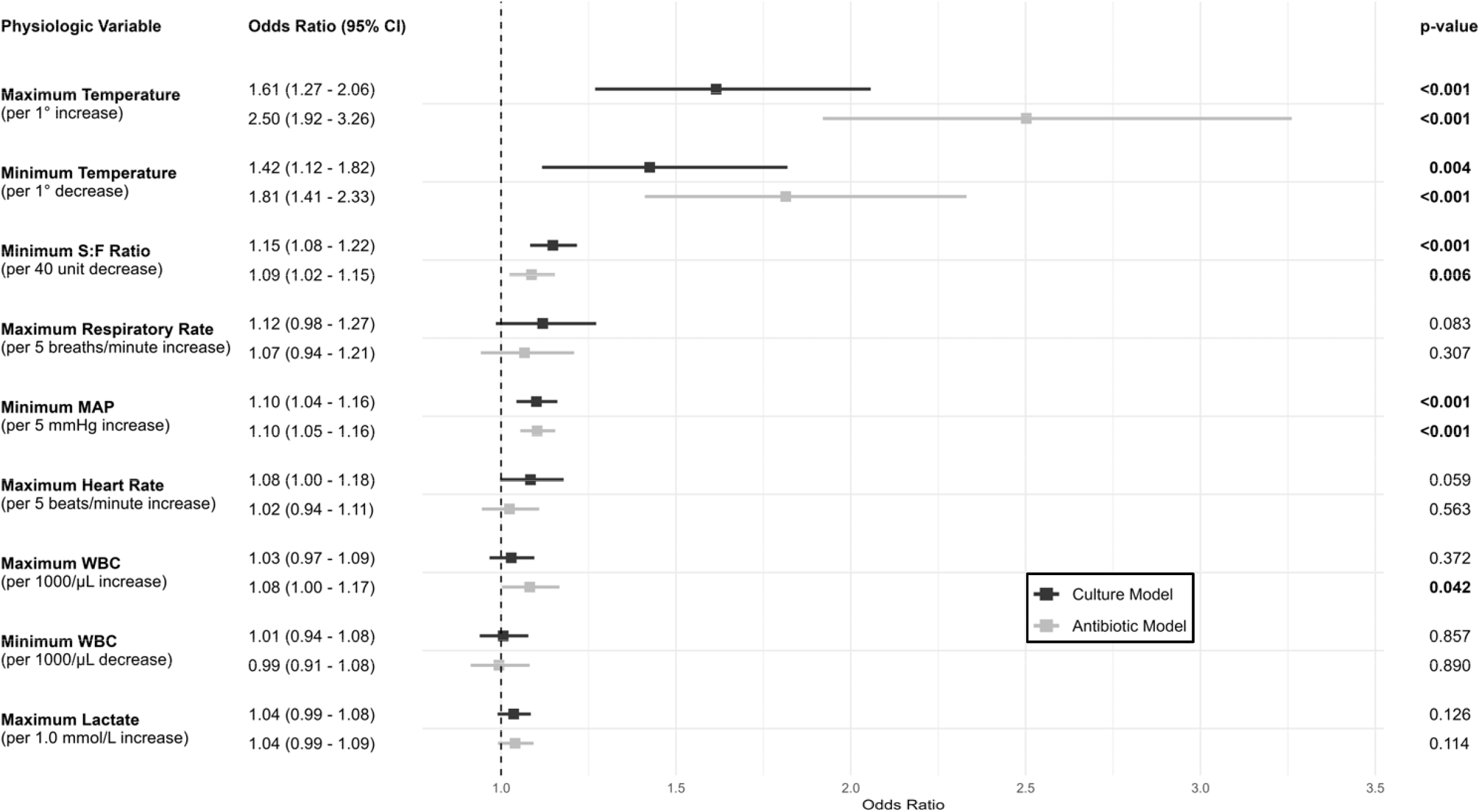
Multivariable logistic regression for the association of physiologic parameters with culture positivity and receipt of broad-spectrum antibiotics at ICU admission.

### Association with Receipt of Antibiotics on ICU Admission

Among all encounters, patients received broad spectrum antibiotics at ICU admission in 44.7% (n=635) of encounters (**Table 2**). This included 76.8% (n=235) of the 306 encounters with positive cultures, 50.4% (n=306) of the 607 encounters with negative cultures, and 18.6% (n=94) of the 506 encounters without any eligible cultures collected during follow-up. The odds of receiving broad-spectrum antibiotics were concordant with the odds of developing culture positivity for all measured physiologic parameters except leukocytosis which was associated with antibiotics receipt but not with culture positivity (**Figure 2**). These results were unaffected by excluding potential contaminant culture results (supplementary **eTable 7**) or encounters without any cultures collected during follow-up (supplementary **eTable 8**) in separate sensitivity analyses.

**Table 2.**
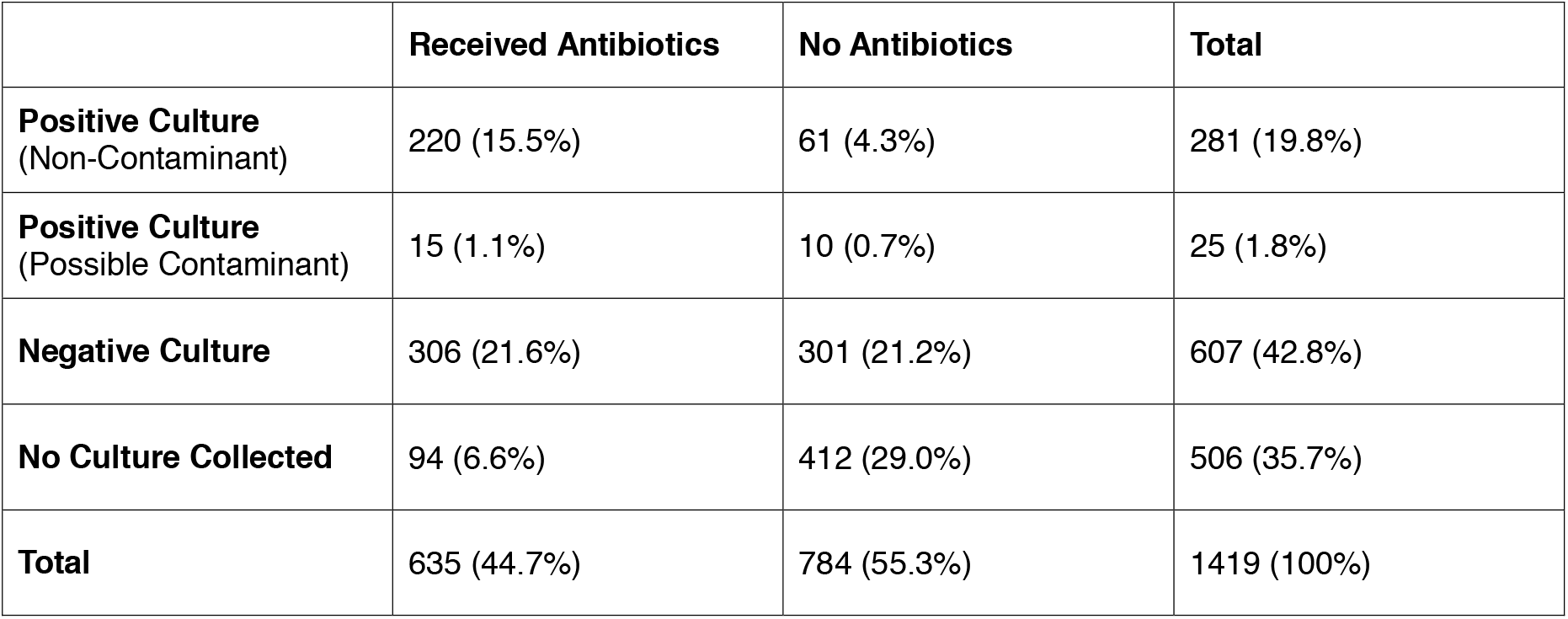
Contingency table of culture positivity and antibiotic receipt status among all eligible ICU encounters.

### Association with Positive Cultures without Antibiotic Receipt

Among the 306 encounters with positive cultures, patients were afebrile (maximal temperature <38 °C) and had a normal WBC count (4,000/μL – 12,000 μL) in 81.7% (n=250) and 48.4% (n=148) of encounters respectively (**Table 3**). In a post-hoc multivariable analysis of these 306 encounters, patients who were afebrile or those with normal WBC were less likely to get antibiotics (**Figure 3**). Among the 71 encounters in which patients with positive cultures did not receive antibiotics at ICU admission (23.2% of all positive cultures), patients were afebrile and had a normal WBC in 97.2% (n=69) and 77.5% (n=55) of encounters respectively.

**Table 3:** Distribution of physiologic parameter measurements among eligible ICU encounters in which patients developed positive cultures at ICU admission.

| Physiologic Parameter<br>(N = 306 per row) | Sub-Group<br>Parameter Value | Positive Cultures<br>N (%) |
| --- | --- | --- |
| Temperature (°C) | > 38 | 56 (18.3) |
|  | 36 – 38 | 166 (54.2) |
|  | < 36 | 84 (27.5) |
| SpO<sub>2</sub>:F<sub>i</sub>O<sub>2</sub> Ratio | ≥ 315 | 214 (69.9) |
|  | < 315 | 92 (30.1) |
| Respiratory Rate (breaths/minute) | > 20 | 200 (65.4) |
|  | ≤ 20 | 106 (34.6) |
| Mean Arterial Pressure (mmHg) | ≥ 65 | 219 (71.6) |
|  | < 65 | 87 (28.4) |
| Heart Rate (beats/minute) | > 90 | 247 (80.7) |
|  | ≤ 90 | 59 (19.3) |
| White Blood Cell Count (count/μL) | > 12,000 | 129 (42.2) |
|  | 4,000 – 12,000 | 148 (48.4) |
|  | < 4,000 | 29 (9.5) |
| Lactate (mmol/L) | > 4 | 201 (65.7) |
|  | 2 – 4 | 79 (25.8) |
|  | < 2 | 26 (8.5) |

**Figure 3:**
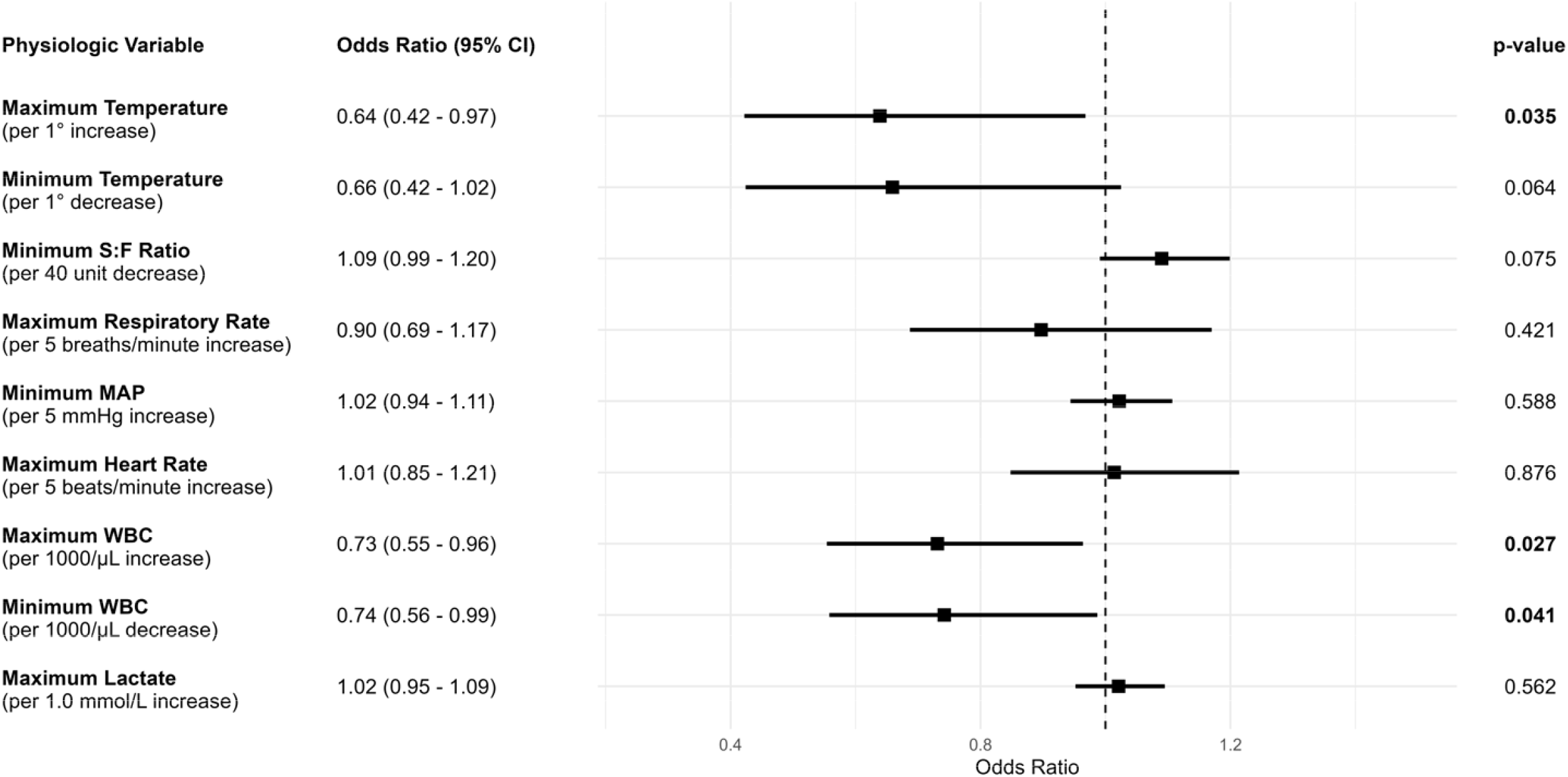
Multivariable logistic regression of the association of physiologic parameters with the odds of antibiotic omission among the n=306 encounters in which patients developed positive cultures at ICU admission.

Among all eligible encounters, temperature and WBC respectively had a bivariate AUROC of only 0.64 and 0.62 for discrimination between patients with and without infection (supplementary **eFigure 1** and see also **Figure 2**).

## DISCUSSION

In this retrospective cohort study of critically ill patients with cirrhosis admitted to the ICU, we found that some—but not all—common physiologic parameters measured at ICU admission were associated with both culture positivity and receipt of broad-spectrum antibiotics. Patients with leukocytosis were more likely to be given antibiotics than those without leukocytosis, but leukocytosis did not differentiate between those with and without positive cultures. Notably, other SIRS criteria—tachycardia, tachypnea, and leukopenia—and elevations in lactate did not differentiate those patients with and without culture-positive infections.

Our findings also highlight that the epidemiology of critically ill patients with cirrhosis may differ from the general cirrhosis population. While spontaneous bacterial peritonitis represents the most common infection in the overall population of patients with cirrhosis, over half of positive cultures in our analysis were from blood.^3^ Additionally, there was a near even split between gram-positive and gram-negative bacterial organisms among positive cultures. These findings were unchanged in a sensitivity analysis after excluding potential contaminant culture results and support the use of broad-spectrum antibiotics to cover both gram-positive and gram-negative organisms in critically ill patients with cirrhosis presenting with potential sepsis at the time of ICU admission.

Our data allow us to only speculate on the failures of certain classic signs of infection to differentiate among cirrhotic patients with and without infection. Patients with cirrhosis not only experience baseline tachycardia in the setting of hyperdynamic circulation, but they may be unable to further augment their heart rate in the setting of infections due to beta blocker therapy ^4^ or variable response to volume expansion.^13^ Increases in lactate and respiratory rate have been shown to shorten time to antibiotics among unselected patients hospitalized with sepsis, but not in our analysis.^41^ Baseline lactic acidosis and tachypnea in patients with cirrhosis may not only preclude the detection of associations with culture positivity, but also lead clinicians to disregard these potential warning signs in this population.^4^

Temperature and WBC poorly discriminated the risk of infection in these patients in both bivariate and multivariable analyses. The presence of hyperthermia and WBC abnormalities has been shown to improve the timely administration of antibiotics in the general sepsis literature more than the presence of shock.^41^ Unfortunately, fever and abnormal WBC may be absent in critically ill patients with cirrhosis presenting with infection.^3^ Indeed, among encounters with positive cultures in our analysis, the majority of pateints (81.7%) were afebrile and half (48.4%) had normal WBC. In a post-hoc sensitivity analysis of these encounters with positive cultures, we further found that the lack of hyperthermia and abnormal WBC was associated with the omission of broad-spectrum antibiotics at ICU admission. Among encounters in which patients with positive culture did not receive antibiotics, nearly all patients (97.2%) were afebrile and most (77.5%) had a normal WBC. Given the severe morbidity and mortality associated with delayed treatment of infections in patients with cirrhosis, it is imperative that clinicians do not anchor on WBC and hyperthermia as the sole determinants of antibiotic administration in this population.

To our knowledge, this is the first study investigating the relationship between the bedside physiology of critically ill patients with cirrhosis presenting to the ICU with both (1) the development of new culture-confirmed infections and (2) early empiric broad-spectrum antibiotic receipt at ICU admission. These findings may inform guidelines for the detection and management of infections in this high-risk population that continues to experience disproportionately high morbidity and mortality despite advances in sepsis management.

There are important limitations to this study. While the data is sourced from a diverse 5-hospital health system with both academic and community representation, they may not be generalizable to other systems where culture collection practices and antibiotic use patterns may differ. Additionally, our primary outcome was limited to microbiologically-confirmed culture results to limit potential confounding in the setting of diagnostic uncertainty. There are likely some patients with culture-negative infections that are not accounted for in our study, particularly given the prevalence of culture-negative spontaneous bacterial peritonitis in this population.^42,43^

## CONCLUSION

At the time of ICU admission, the association of traditional physiologic markers of infection with culture positivity may be weak and divergent with antibiotic administration in critically ill patients with cirrhosis. Our findings suggest that patients with culture-positive infections who do not manifest hyperthermia and WBC abnormalities are at increased risk for delayed use of antibiotics.

## Data Availability

All data produced in the present study are available upon reasonable request to the authors.

## ACKNOWLEDGEMENTS

The authors wish to thank Taylor Bernstein for her expert project management.

