## Supplemental Materials for "Characteristics associated with culture-positivity versus antibiotic administration in patients with cirrhosis on ICU admission: a retrospective cohort study in 5 hospitals"

**ACKNOWLEDGEMENTS:**

The authors wish to thank Taylor Bernstein for her expert project management.

**FINANCIAL DISCLOSURE AND CONFLICTS OF INTEREST:**

Dr. Davis is currently receiving a grant (F32HL182325) from NHLBI. Dr. Iwashyna currently leads a grant (R01HL169533) from NHLBI, a grant (R01EY036670) from NEI, and discloses work as a consultant and potential expert witness to Roots Community Health of Oakland, California, on pulse oximetry unrelated to the contents of this paper.

**CORRESPONDING AUTHOR:**

Andrew J. Davis

Johns Hopkins University

1830 E Monument Street, 5th Floor

Baltimore, MD 21287

**eTable 1: Consensus ICD-10 code set used to identify cirrhosis in electronic health record.**

| **ICD-10 Code** | **Description** |
| --- | --- |
| K70.3  K70.30  K70.31 | Alcoholic cirrhosis of liver  Alcoholic cirrhosis of liver without ascites  Alcoholic cirrhosis of liver with ascites |
| K72.9  K72.90  K72.91 | Hepatic failure, unspecified  Hepatic failure, unspecified without coma  Hepatic failure, unspecified with coma |
| K74.6  K74.60  K74.69 | Other and unspecified cirrhosis of liver  Unspecified cirrhosis of liver  Other cirrhosis of liver |
| K76.6 | Portal Hypertension |
| K76.7 | Hepatorenal syndrome |
| I85.0  I85.00  I85.01  I85.10  I85.11 | Esophageal varices  Esophageal varices without bleeding  Esophageal varices with bleeding  Secondary esophageal varices without bleeding  Secondary esophageal varices with bleeding |
| I85.9 | Oesophageal varices without bleeding |
| I98.2 | Esophageal varices without bleeding classified everywhere |
| I98.3 | Esophageal varices with bleeding in disease classified elsewhere |
| Hospital encounters during which patients were admitted to an intensive care unit within the studied health system during the enrollment period were screened for inclusion if the specific hospital encounter was associated with at least one of the ICD-10 codes contained in the table above as per the consensus code set developed by Shearer et al 2022.^†^  ^†^Shearer, J. E. et al. Systematic review: development of a consensus code set to identify cirrhosis in electronic health records. Alimentary Pharmacology & Therapeutics 55, 645-657 (2022). <https://doi.org/10.1111/apt.16806> | |

| **eTable 2: Physiologic parameter measurements from all eligible ICU encounters before and after imputation.** | | |
| --- | --- | --- |
| **Characteristic** | **Pre-Imputation** N = 1,419*^1^* | **Post-Imputation** N = 1,419*^1^* |
| Max. Temperature (°C) | 36.8 (36.6, 37.1) | 36.8 (36.6, 37.1) |
| Missing, n (%) | 6 (1.1) | 0 (0) |
| Min. Temperature (°C) | 36.3 (36.0, 36.6) | 36.3 (36.0, 36.6) |
| Missing, n (%) | 21 (1.5) | 0 (0) |
| Max. Heart Rate (bpm^2^) | 103 (87, 119) | 103 (87, 119) |
| Missing, n (%) | 3 (0.2) | 0 (0) |
| Min. MAP^3^ (mmHg) | 82 (68, 97) | 82 (68, 97) |
| Missing, n (%) | 10 (0.7) | 0 (0) |
| Max. Respiratory Rate (bpm^4^) | 20 (18, 27) | 20 (18, 27) |
| Missing, n (%) | 12 (0.8) | 0 (0) |
| Min. S:F Ratio | 452 (429, 462) | 448 (413, 462) |
| Missing, n (%) | 255 (18) | 0 (0) |
| Max. WBC^5^ (count/μL) | 8.6 (5.6, 13.3) | 8.7 (5.9, 12.9) |
| Missing, n (%) | 156 (11) | 0 (0) |
| Min. WBC^5^ (count/μL) | 7.7 (5.0, 12.1) | 7.9 (5.3, 11.8) |
| Missing, n (%) | 156 (11) | 0 (0) |
| Max. Lactate (mmol/L) | 3.9 (2.5, 6.7) | 4.2 (2.9, 6.2) |
| Missing, n (%) | 395 (28) | 0 (0) |
| ^1^n (%); Median (Q1, Q3), ^2^mean arterial pressure, ^3^white blood cell. | | |

**eTable 3: List and count of censored physiologic parameter values.**

| **Physiologic Measure** | **Value (Count)** |
| --- | --- |
| Maximum Temperature | 64.1 (1) |
| Minimum Temperature | 0 (2)  1 (1)  2.6 (1)  2.7 (1)  10 (1) |
| Minimum Mean Arterial Pressure (MAP) | 2 (1)  12.3 (1) |
| Maximum Respiratory Rate | 82 (1)  100 (1)  136 (1)  149 (1) |

**eTable 4: List of culture results considered negative (non-pathogenic).**

| **Culture Source** | **Culture Result** |
| --- | --- |
| **Urine** | >100,000 CFU/mL Candida albicans  >100,000 CFU/mL Candida dubliniensis  >100,000 CFU/mL Candida glabrata  >100,000 CFU/mL Candida krusei  >100,000 CFU/mL Candida tropicalis  60,000 CFU/mL Candida albicans  50,000 CFU/mL Candida glabrata  20,000 CFU/mL Candida albicans  >10,000 CFU/mL Aerococcus sanguinicola  >10,000 CFU/mL Candida albicans  >10,000 CFU/mL Candida krusei  >10,000 CFU/mL Gardnerella vaginalis  <10,000 CFU/mL Klebsiella pneumoniae complex  <10,000 CFU/mL Non-Lactose fermenting, Gram negative bacilli  <10,000 CFU/mL Presumptive Enterococcus species  <10,000 CFU/mL Yeast  Candida albicans  candida albicans  candida glabrata  candida krusei  candida lusitaniae  gardnerella vaginalis  lactobacillus species  yeast  yeast previously reported as staphylococcus species, coagulase negative. |
| **Upper Respiratory** | candida dubliniensis  Light growth Candida glabrata  Light growth Yeast  Light growth Yeast, not Cryptococcus neoformans/gattii  Heavy Growth Yeast, not Cryptococcus neoformans/gattii  Moderate growth Yeast, not Cryptococcus neoformans/gattii  Very Light growth Yeast, not Cryptococcus neoformans/gattii  yeast  Yeast, not Cryptococcus neoformans/gattii  yeast, not cryptococcus neoformans/gattii |

**eTable 5: Distribution of sources for positive cultures before and after excluding potential contaminants.**

| **Culture Sources** | **All Cultures**  N = 306 | **Excluding Potential Contaminants** N = 281 |
| --- | --- | --- |
| Blood | 172 (56.2%) | 144 (51.2%) |
| Genitourinary | 85 (27.8%) | 82 (29.2%) |
| Upper Respiratory Tract | 61 (19.9%) | 61 (21.7%) |
| Ascites | 29 (9.5%) | 27 (9.6%) |
| Bronchoalveolar Lavage | 7 (2.3%) | 7 (2.5%) |
| Intra-abdominal, Non-Ascites | 6 (2.0%) | 5 (1.8%) |
| Cerebrospinal Fluid | 2 (0.7%) | 2 (0.7%) |
| Pleural Cavity | 2 (0.7%) | 2 (0.7%) |
| ^1^n (%). Percentages sum to greater than 100%, as each patients in each encounter could have positive cultures from multiple sources. | | |

**eTable 6: List of potential contaminant culture results excluded in the sensitivity analysis.**

| **Culture Source** | **Culture Result** |
| --- | --- |
| **Blood** | Coag-Negative Staphylococcus (S. epidermidis  staphylococcus species, coagulase negative  Staphylococcus species, coagulase negative  Coag-Negative Staphylococcus (S. hominis)  Cutibacterium (Propionibacterium) acnes  coag-negative staphylococcus ( s. epidermidis)  Coag-Negative Staphylococcus (S. haemolyticus)  Dermabacter hominis  Micrococcus luteus  Staphylococcus epidermidis  coag-negative staphylococcus ( s. hominis)  Coag-Negative Staphylococcus (S. simulans)  Corynebacterium amycolatum  Staphylococcus capitis  Staphylococcus pasteuri |
| **Genitourinary** | 30,000 CFU/mL Coag-Negative Staphylococcus (S. epidermidis)  >100,000 CFU/mL Staphylococcus epidermidis  coag-negative staphylococcus ( s. epidermidis)  staphylococcus epidermidis |
| **Ascites** | Coag-Negative Staphylococcus (S. epidermidis)  staphylococcus species, coagulase negative  Staphylococcus species, coagulase negative  coag-negative staphylococcus ( s. hominis) |
| **Intra-abdominal,**  **Non-Ascites** | Light growth Bacteroides fragilis  Light growth Bacteroides thetaiotaomicron group  Light growth Enterococcus faecium  Very Light growth Klebsiella oxytoca/Raoultella ornithinolytica |

**eTable 7: Sensitivity analysis; multivariable logistic regression for the odds of culture positivity and antibiotic receipt at ICU admission after excluding potential contaminant culture results.**

| **Physiologic Parameter** | **All Eligible Encounters**  **(N = 1,419)** | | | |
| --- | --- | --- | --- | --- |
|  | **Culture Positivity** | | **Antibiotic Receipt*** | |
|  | **OR (95% CI)** | **p-value** | **OR (95% CI)** | **p-value** |
| **Maximum Temperature**  *(1°C Increase)* | 1.40 (1.10, 1.79) | **0.007** | 2.50 (1.92, 3.26) | **<0.001** |
| **Minimum Temperature**  *(1°C Increase)* | 1.22 (0.96, 1.54) | 0.103 | 1.81 (1.41, 2.33) | **<0.001** |
| **Minimum S:F Ratio**  *(40 unit Decrease)* | 1.14 (1.07, 1.21) | **<0.001** | 1.09 (1.02, 1.15) | **0.006** |
| **Maximum Respiratory Rate**  *(5 breaths/min Increase)* | 1.11 (0.98, 1.27) | 0.108 | 1.07 (0.94, 1.21) | 0.307 |
| **Mean Arterial Pressure**  *(5 mmHg Decrease)* | 1.09 (1.03, 1.15) | **0.006** | 1.10 (1.05, 1.16) | **<0.001** |
| **Maximum Heart Rates**  *(5 beats/min Increase)* | 1.08 (0.98, 1.18) | 0.119 | 1.02 (0.94, 1.11) | 0.563 |
| **Maximum WBC**  *(1,000/μL Increase)* | 1.04 (0.98, 1.11) | 0.182 | 1.08 (1.00, 1.17) | **0.042** |
| **Minimum WBC**  *(1,000/μL Increase)* | 1.04 (0.97, 1.11) | 0.319 | 0.99 (0.91, 1.08) | 0.890 |
| **Maximum Lactate**  *(1.0 mmol/L Increase)* | 1.05 (1.00, 1.10) | 0.052 | 1.04 (0.99, 1.09) | 0.114 |
| *Antibiotic receipt multivariable logistic regression model is unchanged, as it is not affected by excluding potential contaminant culture results from the culture positivity outcome. | | | | |

**eTable 8. Sensitivity analysis; multivariable logistic regression for the odds of culture positivity and antibiotic receipt at ICU admission after excluding encounters without any cultures collected during follow-up.**

| **Physiologic Parameter** | **All Encounters**  **(N = 913)** | | | |
| --- | --- | --- | --- | --- |
|  | **Culture Positivity** | | **Antibiotic Receipt** | |
|  | **OR (95% CI)** | **p-value** | **OR (95% CI)** | **p-value** |
| **Maximum Temperature**  *(1°C Increase)* | 1.46 (1.15, 1.87) | **0.002** | 2.43 (1.82, 3.25) | **<0.001** |
| **Minimum Temperature**  *(1°C Increase)* | 1.32 (1.04, 1.69) | **0.025** | 1.74 (1.33, 2.26) | **<0.001** |
| **Minimum S:F Ratio**  *(40 unit Decrease)* | 1.14 (1.07, 1.21) | **<0.001** | 1.08 (1.01, 1.16) | **0.020** |
| **Maximum Respiratory Rate**  *(5 breaths/min Increase)* | 1.16 (1.01, 1.32) | **0.032** | 1.11 (0.96, 1.27) | 0.167 |
| **Mean Arterial Pressure**  *(5 mmHg Decrease)* | 1.08 (1.02, 1.14) | **0.011** | 1.09 (1.04, 1.15) | **0.001** |
| **Maximum Heart Rates**  *(5 beats/min Increase)* | 1.06 (0.97, 1.16) | 0.160 | 1.01 (0.92, 1.11) | 0.850 |
| **Maximum WBC**  *(1,000/μL Increase)* | 1.03 (0.97, 1.09) | 0.328 | 1.11 (1.02, 1.22) | **0.022** |
| **Minimum WBC**  *(1,000/μL Increase)* | 1.02 (0.96, 1.09) | 0.520 | 1.03 (0.94, 1.14) | 0.517 |
| **Maximum Lactate**  *(1.0 mmol/L Increase)* | 1.03 (0.99, 1.08) | 0.151 | 1.03 (0.98, 1.09) | 0.253 |

**eFigure 1: Area under the receiver operating characteristic (AUROC) of physiologic parameters for predicting the presence of culture positivity at ICU admission.**

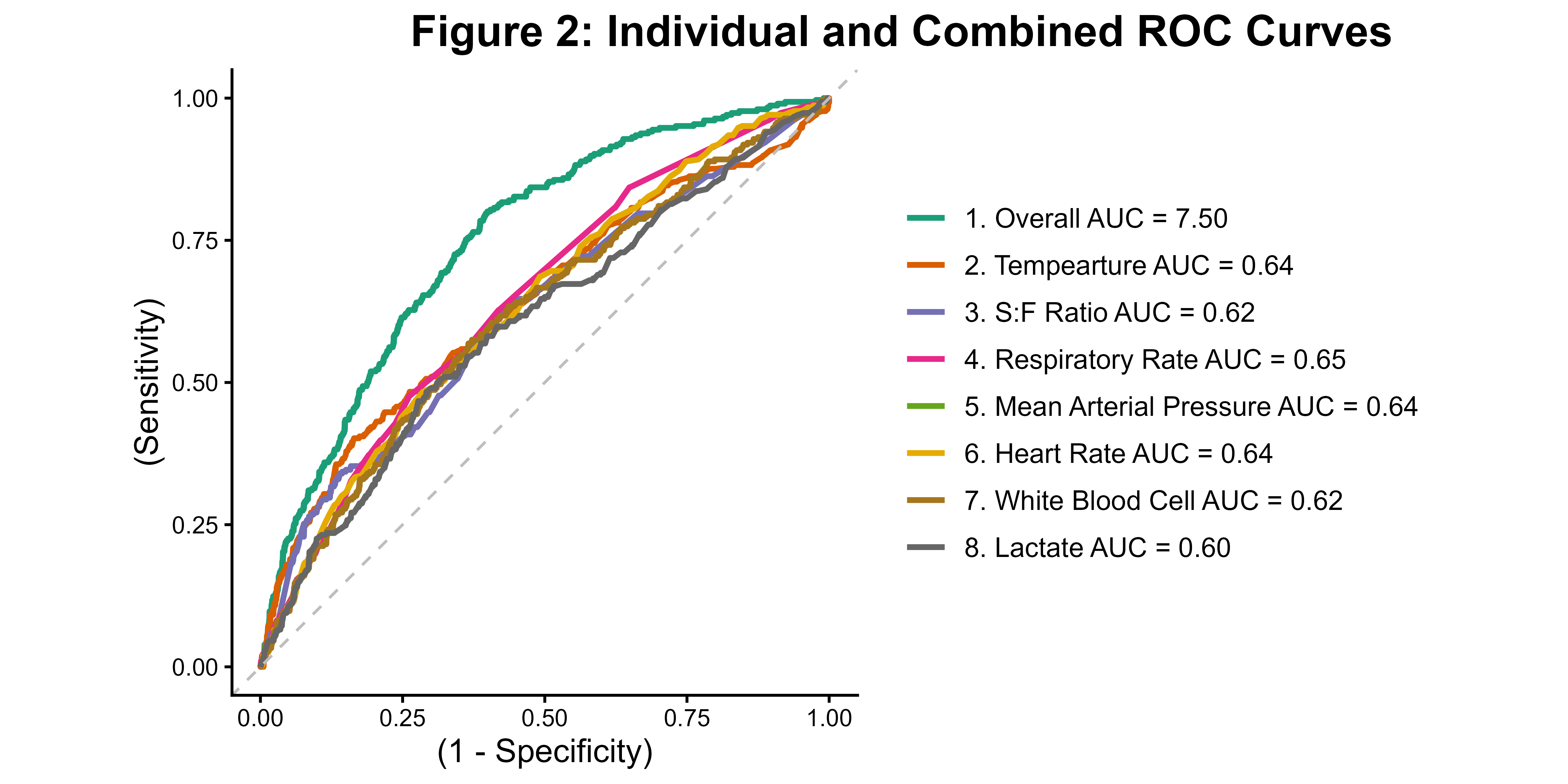

**REFERENCSE**

1	Baudry, T. *et al.* Cirrhotic Patients Admitted to the ICU With Septic Shock: Factors Predicting Short and Long-Term Outcome. *Shock* **52**, 408-413 (2019). <https://doi.org/10.1097/SHK.0000000000001282>

2	Silveira, F. D., Soares, P. H. R., Marchesan, L. Q., Fonseca, R. S. A. D. & Nedel, W. L. Assessing the prognosis of cirrhotic patients in the intensive care unit: What we know and what we need to know better. *World Journal of Hepatology* **13**, 1341-1350 (2021). <https://doi.org/10.4254/wjh.v13.i10.1341>

3	Piano, S., Bunchorntavakul, C., Marciano, S. & Reddy, K. R. Infections in cirrhosis. *The Lancet Gastroenterology & Hepatology* **9**, 745-757 (2024). <https://doi.org/> 10.1016/S2468-1253(24)00078-5

4	Bajaj, J. S., Kamath, P. S. & Reddy, K. R. The Evolving Challenge of Infections in Cirrhosis. *New England Journal of Medicine* **384**, 2317-2330 (2021). [https://doi.org/**10.1056/NEJMra2021808**](https://doi.org/10.1056/NEJMra2021808)

5	Toll, J. F. *et al.* Infections in decompensated cirrhosis: Pathophysiology, management, and research agenda. *Hepatology Communications* **8** (2024). <https://doi.org/> 10.1097/HC9.0000000000000539

6	Albillos, A. *et al.* Cirrhosis-associated immune dysfunction. *Nature Reviews Gastroenterology & Hepatology* **19**, 112-134 (2022). <https://doi.org/10.1038/s41575-021-00520-7>

7	Bruns, T. Risk factors and outcome of bacterial infections in cirrhosis. *World Journal of Gastroenterology* **20**, 2542 (2014). <https://doi.org/10.3748/wjg.v20.i10.2542>

8	Bunchorntavakul, C., Chamroonkul, N. & Chavalitdhamrong, D. Bacterial infections in cirrhosis: A critical review and practical guidance. *World Journal of Hepatology* **8**, 307 (2016). <https://doi.org/10.4254/wjh.v8.i6.307>

9	Simonetto, D. A., Serafim, L. P., Moraes, A. G. D., Gajic, O. & Kamath, P. S. Management of Sepsis in Patients With Cirrhosis: Current Evidence and Practical Approach. *Hepatology* **70**, 418-428 (2019). <https://doi.org/https://doi.org/10.1002/hep.30412>

10	Jimenez, J. V., Garcia-Tsao, G. & Saffo, S. Emerging concepts in the care of patients with cirrhosis and septic shock. *World Journal of Hepatology* **15**, 497-514 (2023). <https://doi.org/10.4254/wjh.v15.i4.497>

11	Fede, G., Privitera, G., Tomaselli, T., Spadaro, L. & Purrello, F. Cardiovascular dysfunction in patients with liver cirrhosis. *Ann Gastroenterol* **28**, 31-40 (2015).

12	Møller, S. & Henriksen, J. H. Cardiovascular complications of cirrhosis. *Gut* **57**, 268-278 (2008). <https://doi.org/10.1136/gut.2006.112177>

13	Durand, F., Kellum, J. A. & Nadim, M. K. Fluid resuscitation in patients with cirrhosis and sepsis: A multidisciplinary perspective. *Journal of Hepatology* **79**, 240-246 (2023). <https://doi.org/https://doi.org/10.1016/j.jhep.2023.02.024>

14	Passino, C. *et al.* Abnormal hyperventilation in patients with hepatic cirrhosis: Role of enhanced chemosensitivity to carbon dioxide. *International Journal of Cardiology* **154**, 22-26 (2012). <https://doi.org/10.1016/j.ijcard.2010.08.066>

15	Benz, F., Mohr, R., Tacke, F. & Roderburg, C. Pulmonary complications in patients with liver cirrhosis. *Journal of Translational Internal Medicine* **8**, 150-158 (2020). <https://doi.org/10.2478/jtim-2020-0024>

16	Moreau, R., Lee, S. S., Soupison, T., Roche-Sicot, J. & Sicot, C. Abnormal tissue oxygenation in patients with cirrhosis and liver failure. *Journal of Hepatology* **7**, 98-105 (1988). <https://doi.org/10.1016/s0168-8278(88)80512-9>

17	Qamar, A. A. & Grace, N. D. Abnormal Hematological Indices in Cirrhosis. *Canadian Journal of Gastroenterology* **23**, 441-445 (2009). <https://doi.org/10.1155/2009/591317>

18	Dasgupta, A. *et al.* Identifying a unique signature of sepsis in patients with pre-existing cirrhosis. *Critical Care* **29** (2025). <https://doi.org/10.1186/s13054-025-05423-6>

19	Pickens, C. I. & Wunderink, R. G. Principles and Practice of Antibiotic Stewardship in the ICU. *Chest* **156**, 163-171 (2019). <https://doi.org/10.1016/j.chest.2019.01.013>

20	Paintsil, E. K., Adu-Asiamah, C. K., Kronsten, V. T., Ntuli, Y. & Shawcross, D. L. Global Trends in Antimicrobial Resistance Among Cirrhosis Patients With Bacteremia: A Systematic Review and Meta-analysis. *Clinical Gastroenterology and Hepatology* **24**, 69-80 (2026). <https://doi.org/10.1016/j.cgh.2025.07.014>

21	Arabi, Y. M. *et al.* Antimicrobial therapeutic determinants of outcomes from septic shock among patients with cirrhosis. *Hepatology* **56**, 2305-2315 (2012). <https://doi.org/10.1002/hep.25931>

22	Prescott, H. C. & Iwashyna, T. J. Improving Sepsis Treatment by Embracing Diagnostic Uncertainty. *Annals of the American Thoracic Society* **16**, 426-429 (2019). <https://doi.org/10.1513/AnnalsATS.201809-646PS>

23	Prescott, H. C. *et al.* Temporal Trends in Antimicrobial Prescribing During Hospitalization for Potential Infection and Sepsis. *JAMA Internal Medicine* **182**, 805 (2022). <https://doi.org/:10.1001/jamainternmed.2022.2291>

24	Gando, S. *et al.* The SIRS criteria have better performance for predicting infection than qSOFA scores in the emergency department. *Scientific Reports* **10** (2020). <https://doi.org/10.1038/s41598-020-64314-8>

25	Marik, P. E. & Taeb, A. M. SIRS, qSOFA and new sepsis definition. *Journal of Thoracic Disease* **9**, 943-945 (2017). <https://doi.org/10.21037/jtd.2017.03.125>

26	Williams, J. M. *et al.* Systemic Inflammatory Response Syndrome, Quick Sequential Organ Function Assessment, and Organ Dysfunction. *CHEST* **151**, 586-596 (2017). <https://doi.org/10.1016/j.chest.2016.10.057>

27	Singer, M. *et al.* The Third International Consensus Definitions for Sepsis and Septic Shock (Sepsis-3). *JAMA* **315**, 801 (2016). <https://doi.org/10.1001/jama.2016.0287>

28	Raith, E. P. *et al.* Prognostic Accuracy of the SOFA Score, SIRS Criteria, and qSOFA Score for In-Hospital Mortality Among Adults With Suspected Infection Admitted to the Intensive Care Unit. *JAMA* **317**, 290 (2017). <https://doi.org/10.1001/jama.2016.20328>

29	Shearer, J. E. *et al.* Systematic review: development of a consensus code set to identify cirrhosis in electronic health records. *Alimentary Pharmacology & Therapeutics* **55**, 645-657 (2022). <https://doi.org/10.1111/apt.16806>

30	Quan, H. *et al.* Coding Algorithms for Defining Comorbidities in ICD-9-CM and ICD-10 Administrative Data. *Medical Care* **43**, 1130-1139 (2005). <https://doi.org/10.1097/01.mlr.0000182534.19832.83>

31	Rice, T. W. *et al.* Comparison of the Sp o 2 /F io 2 Ratio and the Pa o 2 /F io 2 Ratio in Patients With Acute Lung Injury or ARDS. *Chest* **132**, 410-417 (2007). <https://doi.org/10.1378/chest.07-0617>

32	Buuren, S. V. & Groothuis-Oudshoorn, K. mice: Multivariate Imputation by Chained Equations inR. *Journal of Statistical Software* **45** (2011). <https://doi.org/10.18637/jss.v045.i03>

33	Wolk, D. M. *et al.* The American Society for Microbiology’s evidence-based laboratory medicine practice guidelines for the diagnosis of bloodstream infections using rapid tests: a systematic review and meta-analysis. *Clinical Microbiology Reviews* **38** (2025). <https://doi.org/10.1128/cmr.00137-24>

34	Pendleton, K. M., Huffnagle, G. B. & Dickson, R. P. The significance of Candida in the human respiratory tract: our evolving understanding. *Pathogens and Disease* **75** (2017). <https://doi.org/10.1093/femspd/ftx029>

35	Xie, Y. *et al.* Bacterial distributions and prognosis of bloodstream infections in patients with liver cirrhosis. *Scientific Reports* **7** (2017). <https://doi.org/10.1038/s41598-017-11587-1>

36	Yadav, K. & Lewis, R. J. Immortal Time Bias in Observational Studies. *JAMA* **325**, 686 (2021). <https://doi.org/10.1001/jama.2020.9151>

37	Bus, L. D., Arvaniti, K. & Sjövall, F. Empirical antimicrobials in the intensive care unit. *Intensive Care Medicine* **50**, 1338-1341 (2024). <https://doi.org/10.1007/s00134-024-07453-0>

38	Park, J. G. Decompensated cirrhosis and antibiotic prophylaxis: striking a delicate balance. *The Korean Journal of Internal Medicine* **39**, 369-370 (2024). <https://doi.org/10.3904/kjim.2024.119>

39	Zeileis, A., Köll, S. & Graham, N. Various Versatile Variances: An Object-Oriented Implementation of Clustered Covariances in R. *Journal of Statistical Software* **95** (2020). <https://doi.org/10.18637/jss.v095.i01>

40	R: A Language and Environment for Statistical Computing (R Foundation for Statistical Computing, Vienna, Austria, 2021).

41	Wayne, M. T. *et al.* Temporal Trends and Hospital Variation in Time-to-Antibiotics Among Veterans Hospitalized With Sepsis. *JAMA Network Open* **4**, e2123950 (2021). <https://doi.org/10.1001/jamanetworkopen.2021.23950>

42	Kamani, L., Mumtaz, K., Ahmed, U. S., Ali, A. W. & Jafri, W. Outcomes in culture positive and culture negative ascitic fluid infection in patients with viral cirrhosis: cohort study. *BMC Gastroenterology* **8**, 59 (2008). <https://doi.org/10.1186/1471-230X-8-59>

43	Santoiemma, P. P., Dakwar, O. & Angarone, M. P. A retrospective analysis of cases of Spontaneous Bacterial Peritonitis in cirrhosis patients. *PLOS ONE* **15**, e0239470 (2020). <https://doi.org/10.1371/journal.pone.0239470>
